# Modified Long-Axis In-Plane Ultrasound Versus Short-Axis Out-of-Plane Ultrasound For Radial Arterial Cannulation: A Prospective Randomized Controlled Trial

**DOI:** 10.1101/19005496

**Authors:** Jiebo Wang, Liangcheng Zhang, Qijian Huang, Zhongmeng Lai, Guohua Wu, Lei Lin, Junle Liu, Xianfeng Weng

**Author notes:** This author contributed equally to this work and should be considered as a cocorresponding author.

## Abstract

**BACKGROUND:** Given a low first-pass success rate of the conventional SAX (short-axis) or LAX (long-axis) approach, ultrasound-guided radial artery cannulation in adults with radial artery diameter less than 2.2 mm may be still challenging.

**OBJECTIVE:** To assess the efficacy of modified long-axis in-plane(M-LAIP) versus short-axis out-of-plane (SAOP) or conventional palpation(C-P) approaches for ultrasound-guided radial artery cannulation.

**DESIGN:** A prospective, randomized and controlled trial.

**SETTING:** Operating room in a tertiary university hospital, from 1 July 2018 to 24 November 2018.

**PATIENTS:** A total of 201 patients (age 18 to 85 years, the diameter of the radial artery less 2.2 mm) were included. Patients with history of forearm surgery, ulnar artery occlusion, abnormal Allen test, etc, were excluded from this study.

**INTERVENTIONS:** All patients were randomized 1:1:1 to M-LAIP, SAOP or C-P.

**MAIN OUTCOME MEASURES:** The primary outcome was the cannulation success rate. Secondary outcomes included first location time and cannulation time, number of attempts.

**RESULTS:** The cannulation success rate was significantly higher in the M-LAIP group than in the SAOP group or C-P group (first success rate: 80.3% vs 53.8% or 33.8%; p =0.000; total success rate: 93.9% vs 78.5% or 50.8%; p =0.000). First location time (s) was significantly longer in the M-LAIP group compared with the SAOP group (31(28-35[12-44]) vs 15(14-17[10-21]); p =0.000) and the C-P group (31(28-35[12-44]) vs 12(8-13.5 [6-37]); p =0.000). However, the time of cannulation in the M-LAIP group (29(24-45[16-313])) was significantly shorter than that in the SAOP group (45(28.5-135.5[14-346]), p =0.002) and in the C-P group(138(27-308[12-363]), p =0.000). The number of attempts decreased in the M-LAIP group compared with SAOP or C-P group (1.29±0.63 vs 1.8±0.89 or 2.22±0.93, p =0.000).

**CONCLUSION:** The M-LAIP procedure for ultrasound-guided radial artery cannulation can offer a higher success rate of the first-attempt and total cannulation, fewer attempts and less time of cannulation.

**TRIAL REGISTRATION:** The study was registered at ClinicalTrials.gov (http://www.chictr.org.cn/index.aspx, number: ChiCTR-IOR-17011474).

## Introduce

Radial arterial cannulation is a common and frequent invasive procedure for continuous arterial pressure monitoring and arterial blood sampling in operating rooms, intensive care units and the emergency departments.^1, 2^ However, especially for difficult case, such as small artery, the multiple attempts at ultrasound-guided or palpation for radial artery catheterization may cause serious complications, including hemorrhage, hematoma, vasospasm and posterior wall damage.^1-5^

Recently, ultrasound-guided radial artery catheterization has become a research hotspot for its visualization of the targeted artery.^1^ Many studies have shown that ultrasound-guided radial artery cannulation is an effective and safe technique, which may increase the first attempt success rate in small children and adults.^2, 6^

The structures of radial artery may be viewed with ultrasound in three orientations: the short-axis (SAX), the long-axis (LAX) and the oblique-axis (OAX).^7^ Nevertheless, there is insufficient evidence to definitively support LAX, SAX or OAX in patients undergoing ultrasound-guided radial artery cannulation.^7, 8^ Despite SAX easily allows visualization of the target artery, it makes more difficult for needle tip visualization and control.^7, 9, 10^ The needle visualization can be optimized with LAX, but it is difficult to determine the central axis of the vessel because of so narrow ultrasonic plane^11^ which may lead to the unclear visualization of vessels. Additionally, ultrasound section-thickness may introduce artifacts, which can lead to dislocation of the needle tip when using LAX ultrasonic guidance.^12^

Several studies have shown that, using the conventional SAX or LAX approach, it has a low first-pass success rate of radial artery cannulation, ranging from 51%--76%.^11, 13-16^ To our knowledge, for adults with radial artery diameter less than 2.2 mm, information on the effect of ultrasound-guided radial artery cannulation is lacking.

Therefore, we designed a randomized controlled trial to evaluate benefit of ultrasound-guided radial artery cannulation, using modified long-axis in-plane(M-LAIP) technique. The primary hypothesis was that the M-LAIP ultrasound-guided technique would provide higher success rate of cannulation, fewer attempts and less time of cannulation as compared with the short-axis out-of-plane (SAOP) and conventional palpation(C-P).

## Methors

### Register

Ethical approval for this study (Ethical Committee 2018YF017-01, http://www.chictr.org.cn/index. aspx) was provided by the Ethical Committee for Clinical Investigations, Fujian Medical University Union Hospital (Chairperson Prof Libin Liu) on 24 June 2018.

### Study design

This study was a prospective, randomized and controlled clinical trial.

### Study Patients

A written informed consent in the study was obtained from each of 201 patients undergoing elective surgery between 1 July, 2018 and 24 November, 2018. Adult patients with radial artery diameter less than 2.2 mm were included in the study. Patients with history of forearm surgery, ulnar artery occlusion, ipsilateral radial cannulation within a week before the procedure, coagulation dysfunction, abnormal Allen test, or skin infection at the puncture site were excluded from the trial.

### Randomization and Blinding

Randomisation generated by a computer was stratified and placed in sealed opaque envelopes to ensure balance between the two operators and each technique. Patients were randomized 1:1:1 to M-LAIP, C-PT or SAOP. The envelope was broken by a trained investigator until after the control measurements had been made.

### Study Interventions

In the pre-anesthesia preparation room, a specialist anesthesiologist who did not perform artery cannulation measured the diameter and depth of the artery, using the caliper tool on the ultrasound machine. The same ultrasound machine (SonoSite M-Turbo Color Doppler Ultrasound Diagnostic instrument, Linear Array probe (L25N/13–6; SonoSite Inc)) was used for all the ultrasound-guided cannulation groups. All cannulations were performed in the operating room using a 22-gauge intravenous catheter (Jelco; Smith Medical International Ltd, Rossendale, UK) by two anesthesiologists who had previously performed more than 160 arterial cannulations each year (including 30 M-LAIP, 30 SAOP, and 100 palpation). All procedures were performed before or after induction of general anesthesia according to operator’s preference, and 2% lidocaine could be used for local anesthesia in the puncture site.

In the M-LAIP group, after obtaining the view of long axis artery, the probe was slided to maximize the artery diameter in the ultrasound image. The needle was inserted at the point at which the centre line of the probe contacted the skin. Guiding by the center line on the probe, the needle should always be adjusted into the ultrasound plane during the procedure. When the needle tip was advanced into the vascular lumen, the tip bevel toward the anterior wall of the artery was turned to the posterior wall by rotation of the needle through 180°.

If the needle tip was shown to be located in the artery lumem in the ultrasound image, but there was no backflow of blood in the hub of needle, we thought that the needle tip may enter the lateral wall of vascular or be close to the outside of the artery wall.(Figure 2) The position of the needle tip should be estimated according to the direction of the needle away from the center line of probe(that meaned the needle was not aligned with the center line), or by observing the sequence in which the needle and radial artery disappeared successively in the ultrasound image as the probe was slided slowly parallel to the artery. The needle was pulled back until its tip was above the artery but under the skin, and then the needle angle was readjusted by gently swinging its rod so that the tip appeared above the middle of the superior wall of the artery and was clearly visible. It was advanced again until the cathete was within the lumen of radial artery and the blood appeared in the hub. Then the needle was slided out of the catheter 1-2 cm with the right index finger and thumb. Next, the catheter with needle was pushed into the lumen of the vessel. At last, the needle was completely pulled out and the catheter was connected to the transducer. More details about this technique had been previously described.^17^

**Figure 1.**
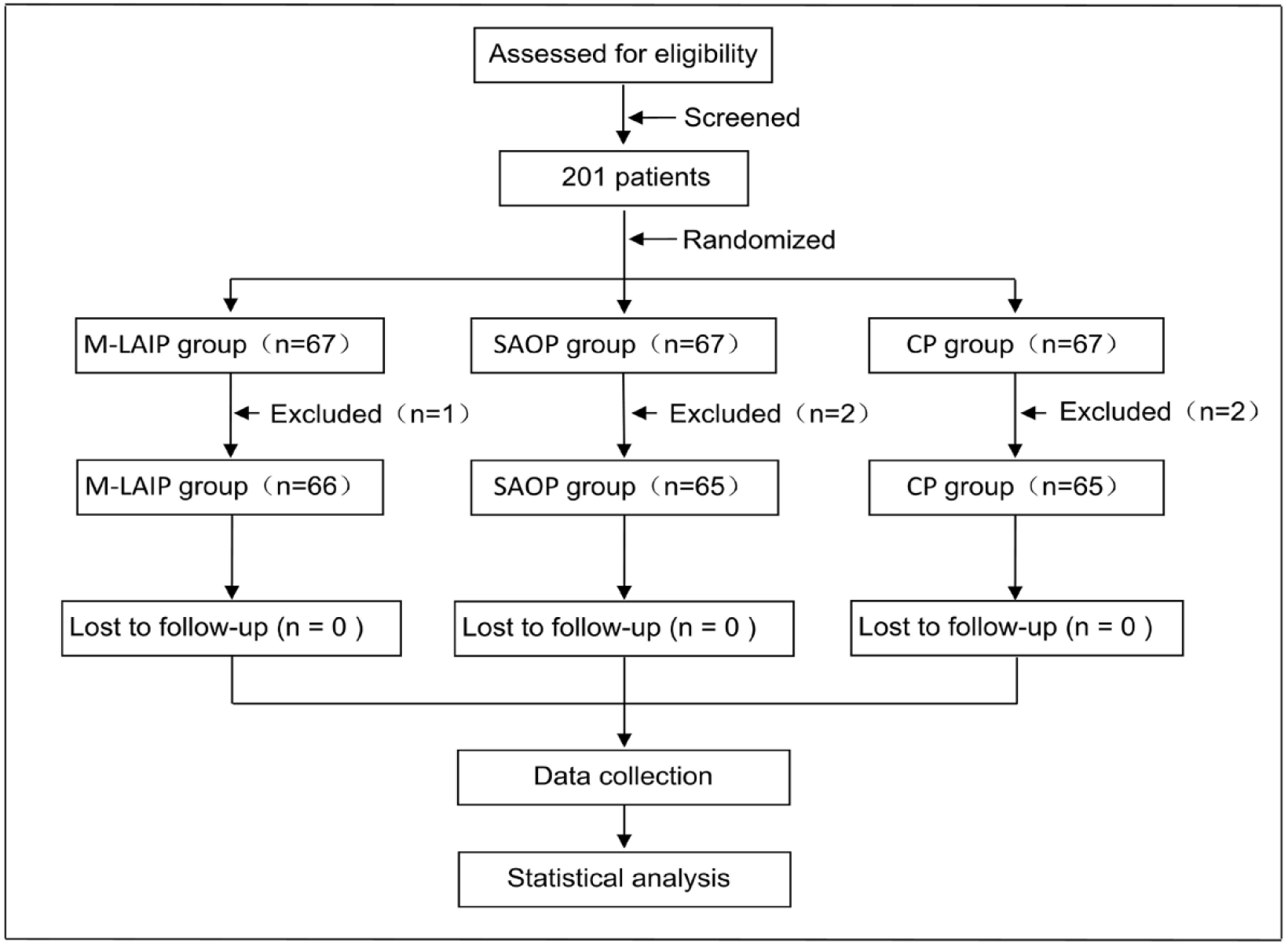
Flow diagram of patient recruitment and randomisation.

**Figure 2.**
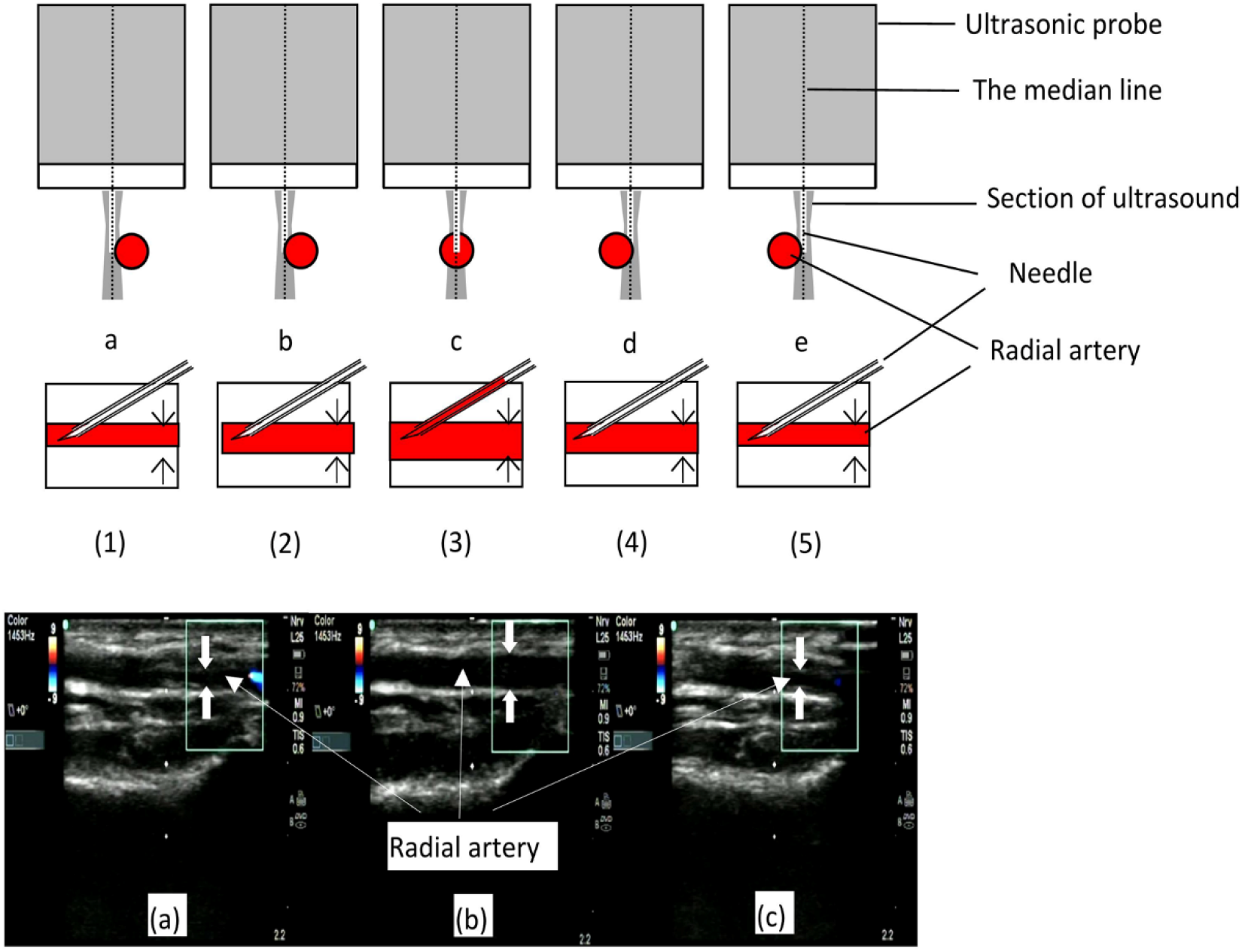
Ultrasonic slice-thickness and diameter of the artery with corresponding images. The slice-thickness of the ultrasonic beam varies with depth (become narrow first, then wider gradually); The diameter of the radial artery varies with the ultrasound probe sliding on the skin and when the central axis of the artery is placed in the ultrasound plane, the diameter of radial artery is maximum((3),(b)).If the ultrasound view of the needle tip inserted into the vascular lumen was obtained, but there is no backflow of blood in the hub((1),(2),(4),(5),(a),(c)), the reason may be that the ultrasonic plane (abcd) deviates from the the central axis of the artery, but part of the vascular lumen is still within ultrasonic beam section, and then the needle tip is inserted into the lateral wall of artery or close to the outside of the vascular wall(a,b,d,e).

In the SAOP group,after viewing the artery in a transverse approach and keeping radial artery in the middle line of the ultrasound screen, the needle was inserted at the puncture point at which the centre line of the probe contacted the skin with an angle of 15°–40° to the skin. Then the needle tip was advanced to the ultrasonic plane by short wiggles. When the needle tip and the catheter was inserted into the arterial lumen and the blood continued to flow into the hub, the catheter was slided into the arterial lumen and attached to the transducer after the needle was removed.

In the palpation group, the operator performed radial artery cannulation by palpating the radial artery pulse, which had been previously described.^17^

### Data Collection

The study data were collected by other anesthesiologists who were not associated with the performance of the procedure. Follow-up was performed by anesthesiologists who were not aware of the grouping on the first and third days after surgery.

### Measurement of Outcomes

The primary outcome was the success of the first and total cannulation. The success of first cannulation was defined as the successful insertion of the cannula with one skin puncture for the first time and the acquisition of arterial waveforms.

Secondary outcomes included cannula insertion failure(considered to be greater than 3 cannulation attempts for a single arm or the attempted cannulation time more than 5 minutes), the number of attempts(defined as the number of skin perforations caused by the needle), the cannulation time (seconds)(defined as the interval between the first skin penetration and confirmation of the arterial waveform on the monitor), first location time (considered to be the interval between the first skin contact of the finger or probe and skin penetration), complications (hematoma thrombosis, oedema or infection), Vasospasm was considered to be any significant resistance during radial artery cannulation manipulation and identified by the operator.^18^ Posterior wall puncture was defined as no backflow of blood after extracting the stylet^16^ and the backflow reoccurred when the catheter was slowly withdrawn.

### Sample size calculation

According to a previous study,^13^ the success rate of cannulation was 51% in the palpation group and 76% in the ultrasound group. With use of the Z-pooled normal approximation method, a sample size of 174 patients (58 per group) provided an 80% power, at a 0.05 two-sided significance level. With an expected 15% loss rate per group, the sample size was finally determined to be 201 patients and 67 patients for each group.

### Statistical Analysis

Statistical analysis was processed using SPSS software (Version 24.0, IBM, USA). The statistics data were expressed as the mean±standard deviation, frequencies (%), the median (interquartile range [IQR]) and odds ratios (95% confidence interval [CI]). The Student’s t-test was used for continuous variables after variables normality and variance homogeneity were tested. The non-parametric tests test was used to compare groups when the normality assumption was not met. The Chi-square (χ^2^) test was used to compare groups for the count data. The 95% confidence intervals were calculated by the Woolf method. It was considered statistically significant for p values <0.05. The Spearman rank correlation coefficien analysis was used for correlations between variables which were not normally distributed.

## Results

In the trial, 201 patients were randomised and five were excluded: cancelled procedures(M-LAIP, n=1, C-PT, n=1), the loss of medical dataset (C-PT, n=1, SAOP, n=1) and transducer faulty for invasive blood pressure (SAOP, n=1), so 196 patients were randomized into the final analysis(Figure 1). General basic characteristics of the patients were summarised in Table 1 and there were no significant differences between study groups.

**Table 1.**
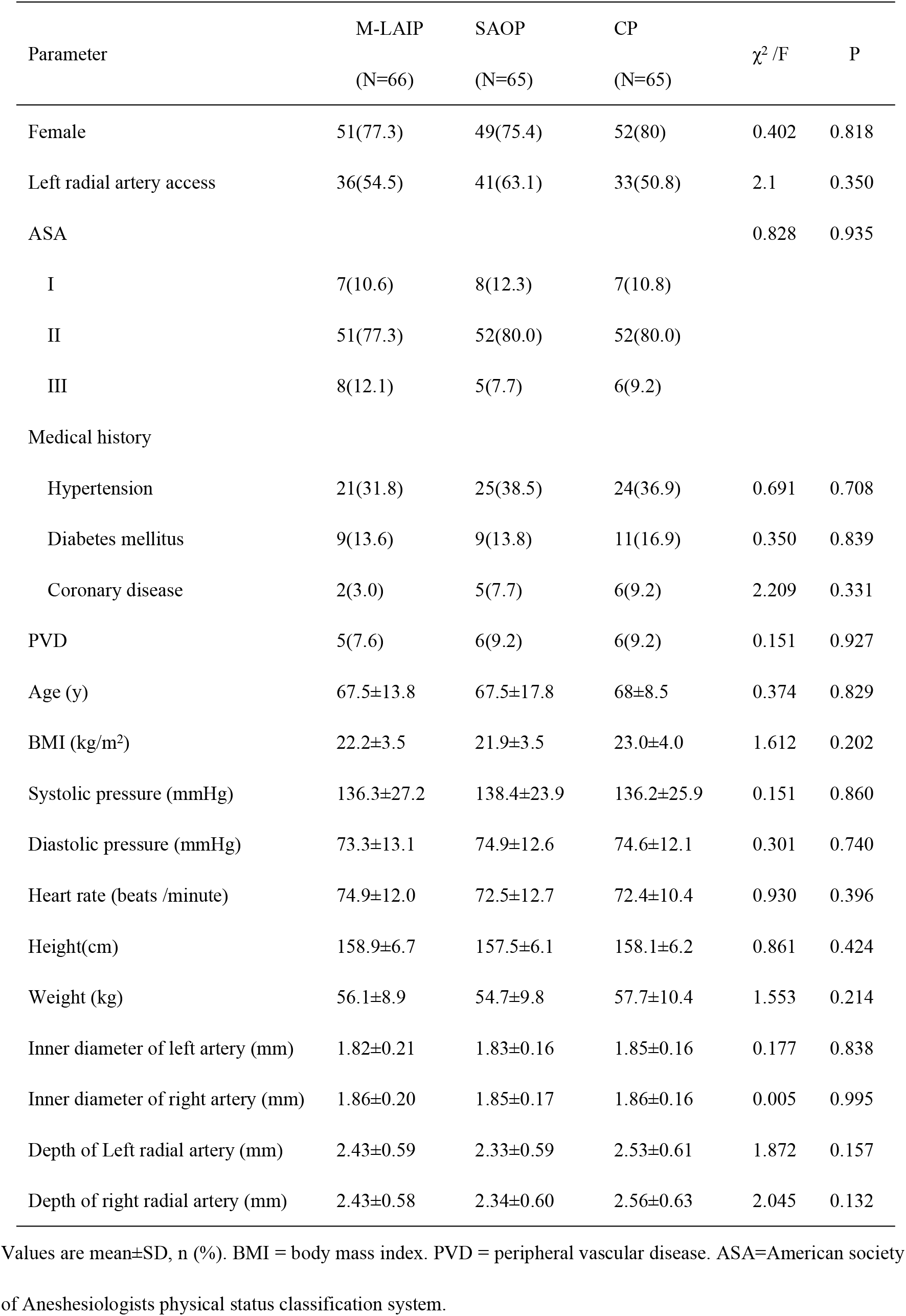
General basic characteristics of the patients.

| Parameter | M-LAIP<br>(N=66) | SAOP<br>(N=65) | CP<br>(N=65) | $\chi^2$ /F | P |
| --- | --- | --- | --- | --- | --- |
| Female | 51(77.3) | 49(75.4) | 52(80) | 0.402 | 0.818 |
| Left radial artery access | 36(54.5) | 41(63.1) | 33(50.8) | 2.1 | 0.350 |
| ASA |  |  |  | 0.828 | 0.935 |
| I | 7(10.6) | 8(12.3) | 7(10.8) |  |  |
| II | 51(77.3) | 52(80.0) | 52(80.0) |  |  |
| III | 8(12.1) | 5(7.7) | 6(9.2) |  |  |
| Medical history |  |  |  |  |  |
| Hypertension | 21(31.8) | 25(38.5) | 24(36.9) | 0.691 | 0.708 |
| Diabetes mellitus | 9(13.6) | 9(13.8) | 11(16.9) | 0.350 | 0.839 |
| Coronary disease | 2(3.0) | 5(7.7) | 6(9.2) | 2.209 | 0.331 |
| PVD | 5(7.6) | 6(9.2) | 6(9.2) | 0.151 | 0.927 |
| Age (y) | 67.5±13.8 | 67.5±17.8 | 68±8.5 | 0.374 | 0.829 |
| BMI (kg/m<sup>2</sup>) | 22.2±3.5 | 21.9±3.5 | 23.0±4.0 | 1.612 | 0.202 |
| Systolic pressure (mmHg) | 136.3±27.2 | 138.4±23.9 | 136.2±25.9 | 0.151 | 0.860 |
| Diastolic pressure (mmHg) | 73.3±13.1 | 74.9±12.6 | 74.6±12.1 | 0.301 | 0.740 |
| Heart rate (beats /minute) | 74.9±12.0 | 72.5±12.7 | 72.4±10.4 | 0.930 | 0.396 |
| Height(cm) | 158.9±6.7 | 157.5±6.1 | 158.1±6.2 | 0.861 | 0.424 |
| Weight (kg) | 56.1±8.9 | 54.7±9.8 | 57.7±10.4 | 1.553 | 0.214 |
| Inner diameter of left artery (mm) | 1.82±0.21 | 1.83±0.16 | 1.85±0.16 | 0.177 | 0.838 |
| Inner diameter of right artery (mm) | 1.86±0.20 | 1.85±0.17 | 1.86±0.16 | 0.005 | 0.995 |
| Depth of Left radial artery (mm) | 2.43±0.59 | 2.33±0.59 | 2.53±0.61 | 1.872 | 0.157 |
| Depth of right radial artery (mm) | 2.43±0.58 | 2.34±0.60 | 2.56±0.63 | 2.045 | 0.132 |

Values are mean±SD, n (%). BMI = body mass index. PVD = peripheral vascular disease. ASA=American society of Anesthesiologists physical status classification system.

The first cannulation success rate was significantly higher in M-LAIP than in both SAOP and C-P (80.3% vs 53.8% and 33.8%; p =0.000; Table 2). Similarly, the total success rate in M-LAIP was 93.9%, which were significantly higher than 78.5% in SAOP or 50.8% in C-PT (p =0.000; Table 2).

**Table 2.**
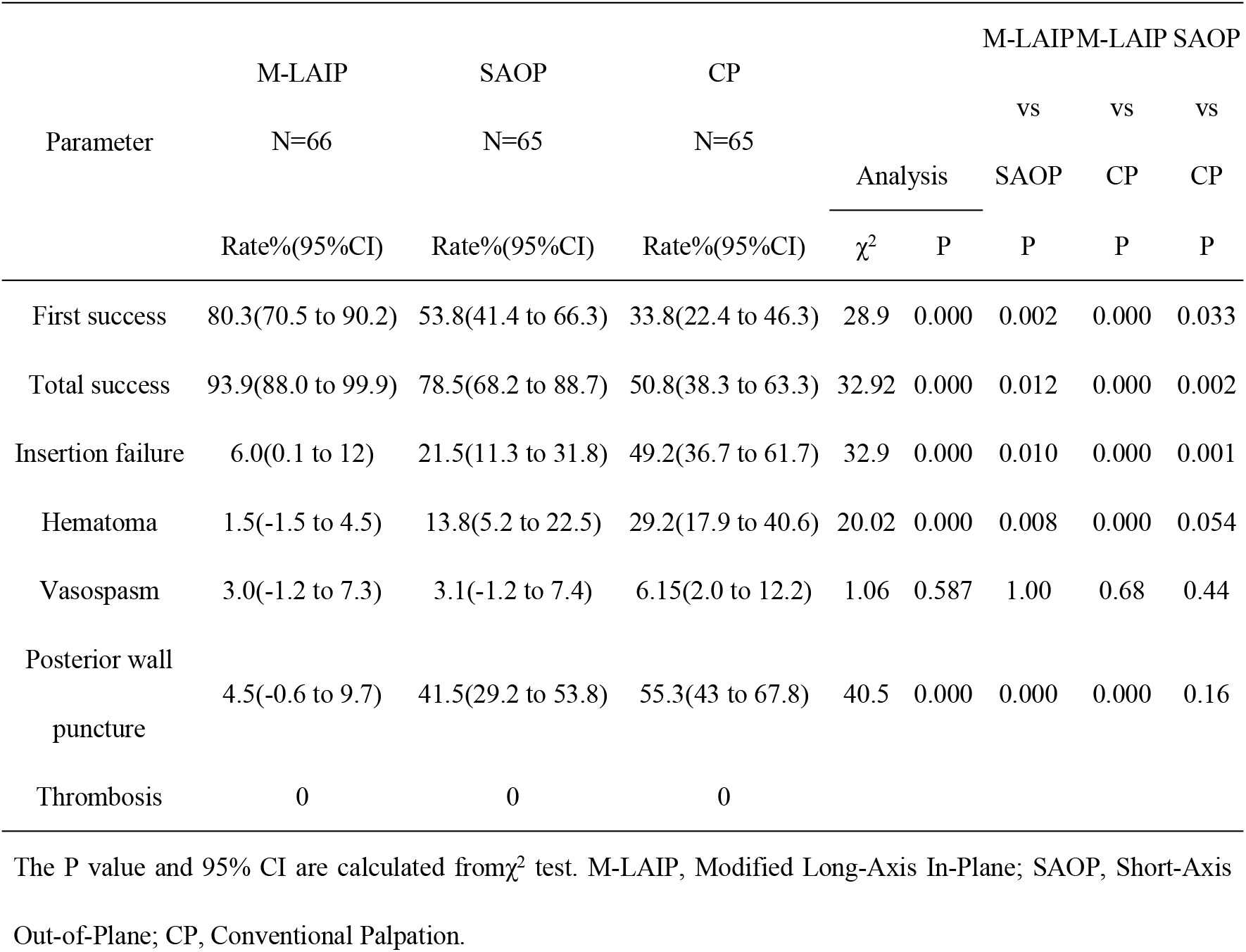
Primary outcomes and complications of radial artery cannulation for three groups.

**Table 3.**
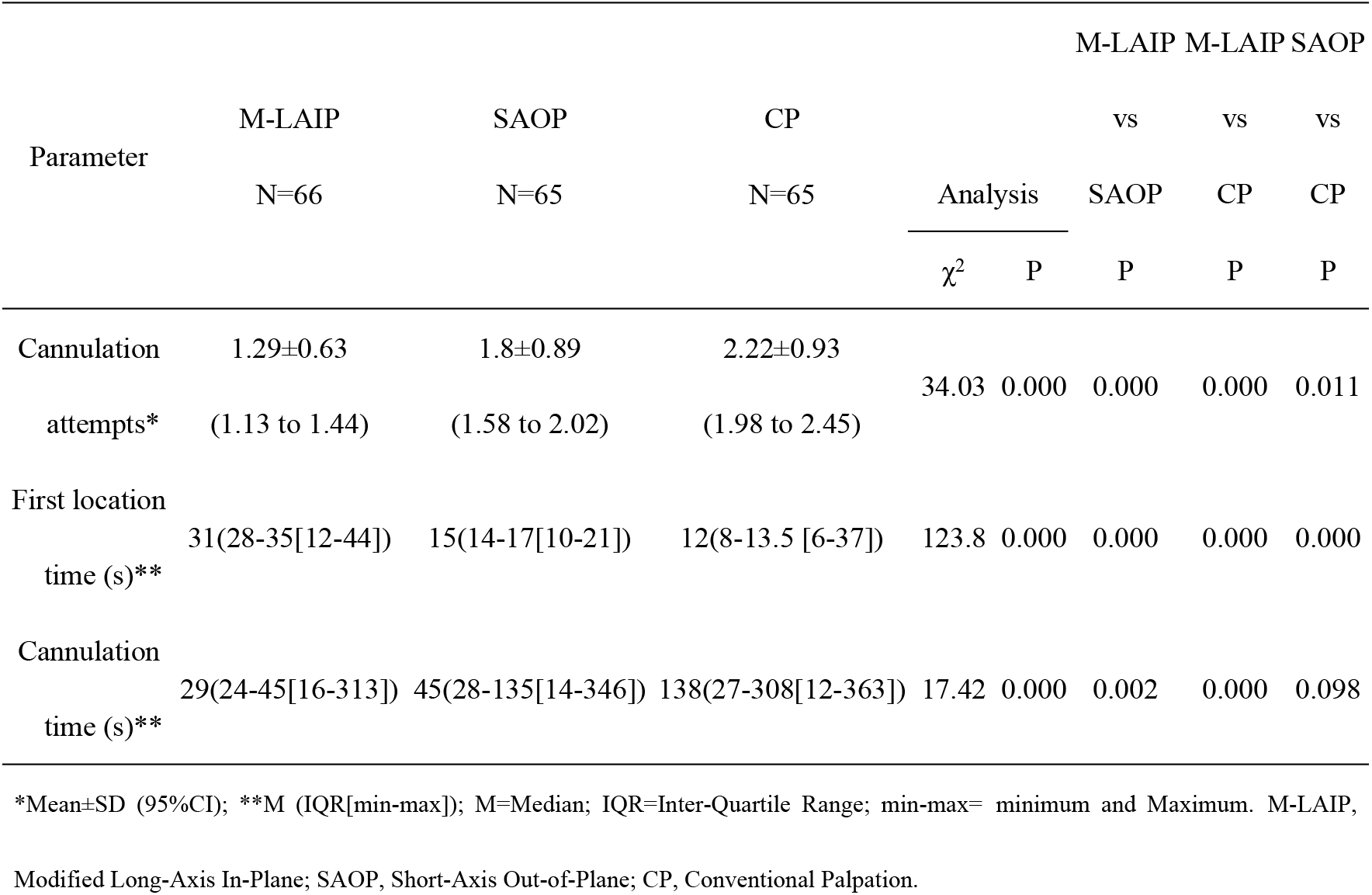
Second outcomes of radial artery cannulation for three groups.

The first and total cannulation success rates were higher in SAOP than in C-PT (53.8% vs 33.8%, p =0.002; 78.5% vs 50.8%, p =0.033, Table 2).

Rate of insertion failure was significantly lower in M-LAIP than in both SAOP and C-PT (6% vs 21.5% and 49.2%; p =0.033, Table 2).

Patients in M-LAIP required fewer attempts than patients in SAOP (1.29±0.63 vs 1.8±0.89, p =0.000, Table 2). Compared with C-P, the number of attempts was fewer in SAOP (1.8±0.89 vs 2.22±0.93, p =0.011, Table 2).

First location time (seconds) was significantly longer in M-LAIP (31(28-35[12-44])) compared with the SAOP ((15(14-17[10-21]); p =0.000) and the C-P (12(8-13.5 [6-37]); p =0.000, table 2). However, the cannulation time (seconds) in M-LAIP (29(24-45[16-313])) was significantly shorter than that in SAOP (45(28.5-135.5[14-346]); p = 0.002) and in C-P (138(27-308[12-363]); p =0.000; Table 2).

The rate of hematoma was significantly lower in M-LAIP than in SAOP (1.5% vs 13.8%, p =0.008, Table 2). However, there were no significant differences between SAOP and C-PT (p =0.054, Table 2).

The rate of vasospasm did not differ significantly in the three groups (p =0.587, Table 2).

The posterior wall puncture rates in SAOP was significantly higher than that in M-LAIP (4.5% vs 41.5%; p =0.000; odds ratio (OR) 14.92; 95% CI, 4.23 to 52.54). However, there were no statistically significant differences between the C-P group and SAOP group (55.3% vs 41.5%, p =0.16). No other severe complications such as thrombosis, edema, infection, were reported.

The Spearman’s rank correlation coefficient analysis revealed a significant but weak correlation between vasospasm and cannulation time (rho =0.279, p =0.000) and number of attempts (rho =0.273, p =0.000). We also found that a significant correlation between hematoma and number of attempts (rho =-0.428, p =0.000).

## Discussion

In this trial we found that M-LAIP approach could increase success rate of the first-attempt and total cannulation and reduce the number of attempts in adults with radial artery diameter less than 2.2 mm as compared with SAOP or C-P approach. We also found that M-LAIP approach may take longer to complete the first location, but it required shorter cannulation time, and the incidence of hematoma complications was lower.

The slice-thickness of ultrasonic beam varies with depth and has a measurable thickness.^19^ The artifacts caused by ultrasound section-thickness will introduce errors of locating the needle tip during the process of ultrasonic guidance.^12^ The ultrasound devices assume that all received echoes come from structures located precisely on the central line of the US beam.^20^ Therefore we define the plane that passes through the center of the ultrasonic section as the ultrasonic plane(abcd), as depicted in Figure 3. When the artery or needle deviates from the ultrasonic plane, without leaving ultrasound section, the ultrasonic view of the artery or needle do not disappear and still be displayed in the ultrasound image (Figure 2). On the contrary, the needle can not be visualized if it is not in the ultrasounic section.^21^ This is the partial ultrasound volume phenomenon caused by the ultrasound section-thickness.

**Figure 3:**
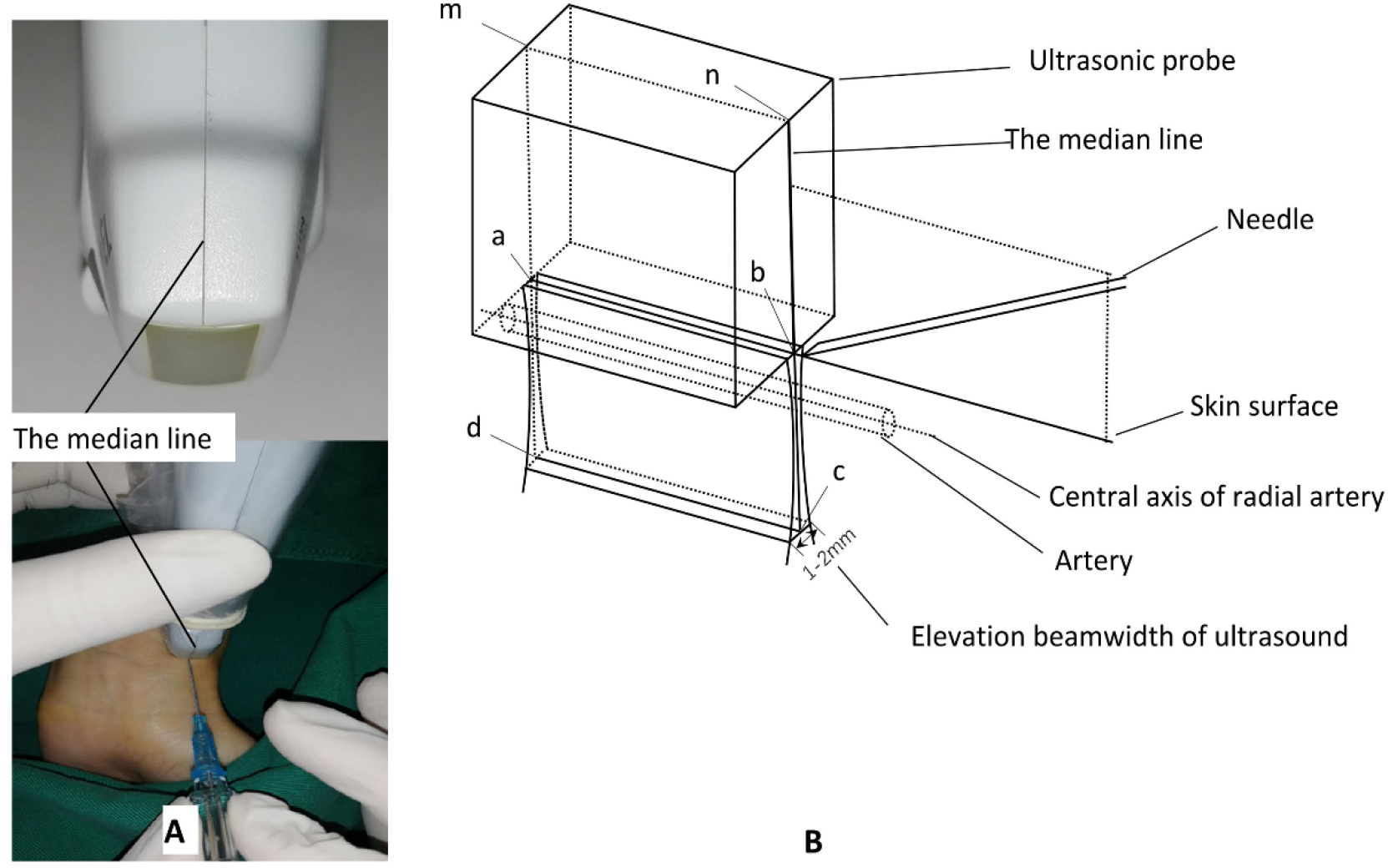
Orientation diagram of Modified long-axis in-plane(M-LAIP) technique. A: The central line of probe is desiged; B: Point a, b, n, m are located in the middle of the ultrasonic probe, respectively. The ultrasonic plane is defined as the plane that passes through the center of the ultrasonic section(abcd); The ultrasounic plane should be aligned with the center axis of radial artery; The needle is adjusted and inserted into the radial artery with the guidance of the central line of probe.

In the long-axis ultrasound view, the diameter of the radial artery varies with the ultrasound probe sliding on the skin, when the central axis of the artery is placed in the ultrasound plane, the maximum diameter of the artery can be obtained (figure 2,c,(3),(b)). Therefore, we have designed a special center line on the side of probe, which is aligned with the ultrasonic plane (figure 3). With the guidance of the central line, the needle is inserted into the ultrasounic plane by adjusting the needle path. (figure 3). As a result, the needle tip aligns with the center of artery.

In practice, we determine the optimal location of the ultrasound probe at the wrist by evaluating the diameter of radial artery (at its maximum diameter) and the signal richness of the blood flow in radial artery in the long-axis ultrasound view (figure 2).^17^ However, the visualization of artery may be unclear sometimes,^7^ which may be associated with the operator’s unfamiliarity with the ultrasound-guided procedures and the different types of ultrasound instruments used. In M-LAIP approach, the special puncture posture and needle-holding mode were adopted,^17^ which have the advantages of stabilizing the hand-held probe, optimizing the visual field, keeping the needle perfectly aligned with the central line of probe, and more precise needle tip control. Our study results also confirm this.

Compared with the SAX approach,^22^ the LAX approach is relatively easy to view the whole needle and the vessel^16^ continuously and the relative position of the needle tip to arteries posterior wall,^13^ which reduces the incidence of posterior wall penetrations and hematoma in our study. The small radial artery (diameter <2.2mm) and the 2-4mm 2-point discrimination limit of finger tip palpation^18^ may be the cause of higher cannulation failure rate, more attempts, longer cannulation time and higher incidence of complications in C-P group. The differences in the first location time in the study groups are in seconds, which may not have much clinical benefit for a procedure.

Berk et al.^13^ supported long axis approach to improve visualization of the needle tip and found that compared with SAX approach, LAX approach had higher first catheter insertion success rates and required fewer attempts and less time. However, the radial artery diameter (4.5mm^2^) in their study might be different from that (1.8mm, 2.54mm^2^) in our study. Additionally, the first cannulation success rate for LAX approach was 76%, which was different from 80.3% in our study.

In-Kyung Song^16^ compared the SAX with LAX approaches to the radial artery cannulation and found that there was no significant difference in the first-pass success rate(68% vs 61.5%) and the total cannulation time in children with artery diameter of 2-2.3mm. That was not consistent with our findings and the difference may be attributable to the different study subjects, ultrasound approach and definition of cannulation success. However, the longer imaging time with the LAX approach was in line with the longer first location time of M-LAIP approach in our study.

Several studys^10,13,16^ show that posterior wall damage rates was higher in SAX than LAX (55.6%-96% for SAX vs 5%-20.4% for LAX), which was in accord with our findings. In SAX method, it is challenging and difficult to identify and track a high echo dot image of needle tip, which may lead to higher posterior wall puncture rate especially for novice ultrasound operators. In our study, the use of the needle tip bevel toward the posterior wall of the artery can further reduce the posterior wall puncture rate.

The research shows the rate of vasospasm occurred rarely (3% to 4%) and there was a correlation between vasospasm and number of attempts or cannulation time,^18^ which was consistent with our findings. Therefore, fewer attempts and shorter cannulation time may reduce the occurrence of spasm.

It had been reported that^11^ incidence of hematoma ranged from 15% to 18% for the ultrasound-guided radial artery cannulation, which was similar to that of SAOP in our study. We found that the increased incidence of hematoma was associated with more attempts, and fewer attempts might induce lower incidence of hematoma in the M-LAIP group.

For cases with difficult cannulation, operator experience in using ultrasound-guided techniques play a key role in successful arterial cannulation, especially for novices who need a procedure with a learning curve.^16, 23^ Therefore, the results of our study were obtained by only two experieced anaesthesiologists.

After being familiar with the use of ultrasound-guided access and imaging,^18^ we believe that the M-LAIP can facilitate clinical work and should be extrapolated to the other area, such as central venous cannulation, nerve block anesthesia, et al.

### Limitations

Two operators were involved in the study, so the Hawthorne effect could not be completely excluded. The absence of statistics on the preparation time of ultrasound probe may negate the average time advantage of cannulation in the ultrasound group. A bias in judgment, recording of procedures, measurement or clinical data cannot be completely ruled out, although the use of an independent observer. Future study is needed regarding infant for cannulation of the radial artery to further evaluate the benefit of the M-LAIP US guidance.

## Conclusion

In conclusion, compared with SAOP and C-P approaches in adults with radial artery diameter less than 2.2 mm, despite longer first location time, use of the M-LAIP technique for arterial catheterization may significantly improve the success rate of the first-attempt and total cannulation, reduce the number of attempts and shorten time of cannulation.

## Data Availability

All data can be searched on Resman clinical trial public manage web.

## Acknowledgements relating to this article

Assistance with the study: We would like to thank Prof Jian zeng (Professor of Anaesthesia in the Department of Anesthesiology at the Fujian Medical University Union Hospital) for his support and quality monitoring of the trial. The authors would like to thank all the nurses, anesthesiologists of the anesthesia operating department.

## Presentation

none.

## Reference

1. Gu WJ, Wu XD, Wang F, Ma ZL, Gu XP. Ultrasound Guidance Facilitates Radial Artery Catheterization: A Meta-analysis With Trial Sequential Analysis of Randomized Controlled Trials. Chest 2016; 149:166–179.

2. Gu WJ, Tie HT, Liu JC, Zeng XT. Efficacy of ultrasound-guided radial artery catheterization: a systematic review and meta-analysis of randomized controlled trials. Critical care (London, England) 2014; 18:R93.

3. Kiberenge RK, Ueda K, Rosauer B. Ultrasound-Guided Dynamic Needle Tip Positioning Technique Versus Palpation Technique for Radial Arterial Cannulation in Adult Surgical Patients: A Randomized Controlled Trial. Anesthesia and analgesia 2018; 126:120–126.

4. Sites BD, Gallagher JD, Cravero J, Lundberg J, Blike G. The learning curve associated with a simulated ultrasound-guided interventional task by inexperienced anesthesia residents. Regional anesthesia and pain medicine 2004; 29:544–548.

5. French JL, Raine-Fenning NJ, Hardman JG, Bedforth NM. Pitfalls of ultrasound guided vascular access: the use of three/four-dimensional ultrasound. Anaesthesia 2008; 63:806–813.

6. Bhattacharjee S, Maitra S, Baidya DK. Comparison between ultrasound guided technique and digital palpation technique for radial artery cannulation in adult patients: An updated meta-analysis of randomized controlled trials. Journal of clinical anesthesia 2018; 47:54–59.

7. Lv Y, Liu H, Yu P, et al. Evaluating the Long-, Short-, and Oblique-Axis Approaches for Ultrasound-Guided Vascular Access Cannulation. Journal of ultrasound in medicine : official journal of the American Institute of Ultrasound in Medicine 2019; 38:347–355.

8. Gao YB, Yan JH, Ma JM, et al. Effects of long axis in-plane vs short axis out-of-plane techniques during ultrasound-guided vascular access. The American journal of emergency medicine 2016; 34:778–783.

9. Batllori M, Urra M, Uriarte E, et al. Randomized comparison of three transducer orientation approaches for ultrasound guided internal jugular venous cannulation. British journal of anaesthesia 2016; 116:370–376.

10. Yoshida T, Nakajima Y, Nakayama Y, et al. Dynamic Needle Tip Positioning for Ultrasound-Guided Arterial Catheterization in Infants and Small Children With Deep Arteries: A Randomized Controlled Trial. Journal of cardiothoracic and vascular anesthesia 2019; 33:1919–1925.

11. Quan Z, Takeshita J, Tian M, Chi P, Cao Y, Li X, Peng K. Modified short-axis out-of-plane ultrasound versus conventional long-axis in-plane ultrasound to guide radial artery cannulation: a randomized controlled trial. Anesthesia and analgesia 2014; 119:163–169.

12. Peikari M, Chen TK, Lasso A, Heffter T, Fichtinger G. Effects of Ultrasound Section-Thickness on Brachytherapy Needle Tip Localization Error, 2011.

13. Berk D, Gurkan Y, Kus A, Ulugol H, Solak M, Toker K. Ultrasound-guided radial arterial cannulation: long axis/in-plane versus short axis/out-of-plane approaches? Journal of clinical monitoring and computing 2013; 27:319–324.

14. Edanaga M, Mimura M, Azumaguchi T, Kimura M, Yamakage M. [Comparison of ultrasound-guided and blindly placed radial artery catheterization]. Masui The Japanese journal of anesthesiology 2012; 61:221–224.

15. Ueda K, Bayman EO, Johnson C, Odum NJ, Lee JJ. A randomised controlled trial of radial artery cannulation guided by Doppler vs. palpation vs. ultrasound. Anaesthesia 2015; 70:1039–1044.

16. Song IK, Choi JY, Lee JH, et al. Short-axis/out-of-plane or long-axis/in-plane ultrasound-guided arterial cannulation in children: A randomised controlled trial. European journal of anaesthesiology 2016; 33:522–527.

17. Wang J, Lai Z, Weng X, et al. Modified Long-Axis In-Plane Ultrasound Technique Versus Conventional Palpation Technique For Radial Arterial Cannulation: A Prospective Randomized Controlled Trial. medRxiv 2019:19001586.

18. Seto AH, Roberts JS, Abu-Fadel MS, et al. Real-time ultrasound guidance facilitates transradial access: RAUST (Radial Artery access with Ultrasound Trial). JACC Cardiovascular interventions 2015; 8:283–291.

19. Goldstein A, Madrazo BL. Slice-thickness artifacts in gray-scale ultrasound. Journal of clinical ultrasound : JCU 1981; 9:365–375.

20. M P, TK C, A L, T H, G F. Effects of ultrasound section-thickness on brachytherapy needle tip localization error. Medical image computing and computer-assisted intervention : MICCAI International Conference on Medical Image Computing and Computer-Assisted Intervention 2011; 14:299–306.

21. Schofer JM, Nomura JT, Bauman MJ, Robert H, Charles S. The “Ski Lift”: A technique to maximize needle visualization with the long-axis approach for ultrasound-guided vascular access. Academic Emergency Medicine Official Journal of the Society for Academic Emergency Medicine 2014; 17:e83–e84.

22. Vogel JA, Haukoos JS, Erickson CL, et al. Is long-axis view superior to short-axis view in ultrasound-guided central venous catheterization? Critical care medicine 2015; 43:832–839.

23. Ueda K, Puangsuvan S, Hove MA, Bayman EO. Ultrasound visual image-guided vs Doppler auditory-assisted radial artery cannulation in infants and small children by non-expert anaesthesiologists: a randomized prospective study. British journal of anaesthesia 2013; 110:281–286.

